# Role of genetics in capturing racial disparities in cardiovascular disease

**DOI:** 10.1101/2023.02.10.23285769

**Authors:** Aritra Bose, Daniel E. Platt, Uri Kartoun, Kenney Ng, Laxmi Parida

## Abstract

The role of race in medical decision-making has been a contentious issue. Insights from history and population genetics suggest considering race as a differentiating marker for medical practices can be influenced by systemic bias, leading to serious errors. This may negatively impact treatment of complex diseases such as cardiovascular disease (CVD). We seek to identify instrumental variables and independently verifiable epidemiological tests of whether diagnoses and treatments impacting severe cardiovascular conditions are racially linked. Using data from the UK Biobank (UKB), we found minimal, non-significant racial differences in log odds ratio (OR) between a range of cardiovascular outcomes such as atrial fibrillation, coronary artery disease, coronary thrombosis, heart failure and cardiac fatality. Genetics classification with respect to principal components vs. racial identification of Black British showed no significant differences in diagnoses or therapeutics for CVD related diseases and their associated comorbidities. However, Black British had significant risk of association with genetically predisposed risk of CVD as captured by polygenic risk scores (PRS) of CVD (OR=1.12; 95%CI:1.034-1.223; *p <* 0.006) as well as in 14 related traits. We used a sub-population based feature selection method to find Townsend Deprivation Index, smoking history, hypertension, PRS for ischemic stroke, low density lipoprotein cholesterol, and type II diabetes as the top features predicting the ethnographic category of Black British with an AUC of 79.5%. Therefore, PRS can be used to understand racial disparities in disease outcome which is otherwise not reflected in clinical factors such as diagnoses outcome status or therapeutics in large observational cohorts such as UKB. PRS yield better predictive power with underrepresented minorities and can improve clinical decision-making.

## Introduction

The field of healthcare is not a stand-alone case of being strife with racial disparities, rather it is a part of a structural racism in our society. *Unequal treatment* [1] was one of the first major reports to state how systemic racism leads to disparities in therapeutics (Rx) and diagnoses (Dx) of common diseases. Even after two decades from that report, we are still dealing with rampant racial disparities in healthcare, recently evident during the COVID-19 pandemic, where Black and Brown communities were disproportionately affected [2]. According to the American Heart Association (AHA), “Race” is considered as a social construct based on phenotype, ethnicity and other indicators of social differentiation resulting in varying access to power and socio-economic resources [3]. Despite mounting evidence that race is not a reliable proxy for genetic difference, use of race-correction algorithms are embedded, often insidiously, within medical practice [4]. Physicians use these race-embedded algorithms to individualize risk assessment and make clinical decisions. The AHA used to assign three additional points to patients identifying as *nonblack* in the Get with the Guidelines-Heart Failure Risk Score which assesses the risk of death in hospitalized patients [5]. But, the reality is often very different from such systemic racism-induced race-embedded risk scores, where in reality African Americans are 30% more likely to die from cardiac heart failure than *nonblack* populations [6]. At the same time, prevalence of coronary artery disease is much lower among black british compared to whites [7, 8] highlighting the complexity of how racial factors impact similar racial classifications in different populations. Similarly, the lack of effectiveness of angiotensin-converting enzyme (ACE) inhibitors in African Americans based on blood pressure measurements was also disputed with conflicting results indicating that they are equally as effective in African Americans compared to Caucasian Americans [9, 10]. Similar examples are ubiquitous in healthcare and clinical decision-making. Hence, alternative approaches such as race-conscious medicines based on genetics are promoted to reduce racial health inequities [11].

Great strides have been made towards understanding race in the context of human genetics since the Human Genome project. Hence, the clinical implementation of self-described racial ethnographic category can often be very different from the population genetics derived notion of “Race” but most race corrections implicitly assume that genetics track reliably with race [4]. While genetic and genomic research is primarily focussed on understanding disease mechanisms and identifying new therapeutic targets, it can also elucidate how genetics can play a significant role in mitigating racial disparities in healthcare and making medicine more equitable. Genomics can capture ancestry in a more precise way, allowing genetic influences to be teased apart from the impact of social and environmental factors [12]. Hence, we can use genetic structure, often represented by principal components (PCs) as a proxy for ethnographic racial categorizations in clinical studies and epidemiology. Another way genetics can aid clinical decision-making is with polygenic risk scores (PRS) which predict complex traits on the basis of genetic data. PRS are poised to improve biomedical outcomes via precision medicine [13] and can be used as an additional biomarker along with the combination of clinical, biochemistry, lifestyle, and historical risk factors [14] which can enhance phenotype prediction accuracy in CVD. Recently, AHA has suggested efficacy, harm and logistics as three criteria to consider for implementation of PRS in cardiovascular care [15]. PRS are often plagued by poor transferability across populations as most large genetic cohorts are overwhelmingly of European descent [14]. Thus, clinical use of PRS may end up exacerbating racial disparities [13].

In this work, we seek to identify instrumental variables and independently verifiable tests of whether Dx and Rx decisions impacting CVD and its comorbidities are racially linked. Specifically, we seek to construct tests to identify whether racial status may contribute to under-diagnosis, under prescribing, and/or increased mortality. It has been shown in prior work that the UK Biobank (UKB) suffers from a well-established “healthy volunteer” effect due to volunteer enrollment in UKB leading to enrollment bias [16]. Black British (BB), who show strong deprivation according to the Townsend deprivation index are most likely impacted by accessibility of treatment which also stems from a mistrust in healthcare due to a history of systemic deprivation [17]. Due to the volunteering nature of enrollment, the distrust in healthcare is mitigated in UKB. The enrollees identified by self-described ethnographic category of BB is 1.54% with 0.21% reported as “mixed”. The genetic history of BB in the UK (3% of entire population) is complex, spanning multiple migrations into Britain, primarily fuelled by colonization of African countries and the West Indies [18]. Therefore, racial classifications can hide distinctive ancestral groups with differential responses to diseases. Understanding any differences in disease impacts should account for possible treatment biases. We sought to track differences along the paths of diagnosis and therapy for CVD related diseases such as Atrial Fibrillation (AF), Coronary Artery Disease (CAD), Coronary Thrombosis (CT), Coronary Fatality (CF), and Heart Failure (HF) as well as their comorbidities such as obesity (OBESE), Chronic Kidney Disease (CKD), diabetes (DIAB), Hyperlipidemia (HL), and Hypertension (HT).

We investigated whether using self-described ethnographic category of BB and other populations in UKB such as East Asians (EAS), South Asians (SAS), mixed and individuals of European ancestry, known hereafter as UK White (UKW) are more susceptible to the above cardiovascular and related diseases in Dx or Rx indicating evidence of racial disparity. We further tested whether alternative genetic measures of ancestrally-correlated indices and polygenic risk scores (PRS) for the above diseases and related clinical variables can be used to capture the racial disparity for BB. We find that PRS for CVD related traits is significantly associated with BB. On the other hand, we either found protective effect or non-significant associations with BB and CVD related Dx or Rx using clinical variables. Hence, the use of genetics in the form of ancestrally informative markers for capturing population structure as a proxy for a racial classification such as BB and PRS in clinical decision-making is an important way to understand racial disparities in large electronic health records (EHR)-linked biobanks. Thus, genetics can play a crucial role in clinical decision-making yielding a more equitable healthcare system.

## 1 Methods

### 1.1 Data

We extracted 104,604 samples with genetic information from UKB along with CVD associated clinical variables related to both Dx and Rx and demographic features such as sex, Townsend Deprivation Index (TDI), and age (we converted the continuous variable representing age to a binary variable *Age65*, representing individuals who are 65 years or older as 1 and rest as 0). We also extracted an enhanced PRS set (ePRS) [19] with PRS of 15 CVD related traits such as AF, CVD, CAD, ischemic stroke (ISS), resting heart rate (RHR), venous thromboembolic disease (VTE), body mass index (BMI), Hypertension (HT), low density lipoprotein (LDL) and high density lipoprotein (HDL) cholesterol levels, etc. A full list of the clinical variables extracted from UKB and the number of people with incident outcome is shown in Figure 1. To identify whether presecriptions were treated differently for BB vs. *nonblack*, we extracted the reported LDL levels just prior to the first prescription of statins (STAT) or fibrates (FIBR). The ICD-9, 10 and other self-reported diagnoses codes used in UK Biobank to extract all clinical information is available in Supplementary Table 1.

**Figure 1:**
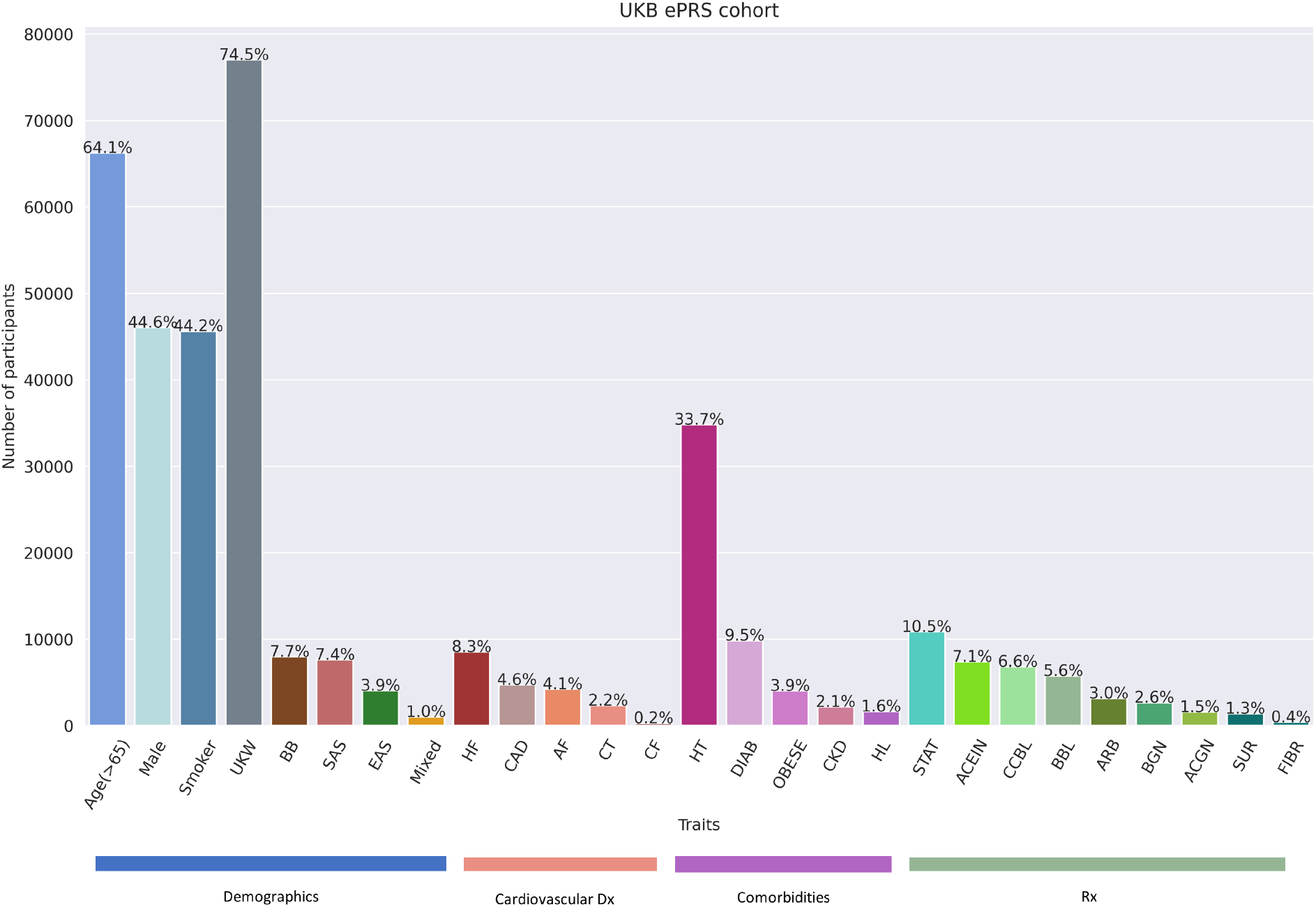
Barplot with the number and proportion of individuals with each trait such as race, demographics, clinical variables, etc. in the UKB ePRS cohort. Variables include (in order as they appear) grouped and colored by their class such as Demographics - Age (*>* 65): individuals older than 65 years; male participants; smoking status; BB: Black British; EAS: East Asians; Mixed: mixed ancestry individuals; SAS: South Asians; UKW: individuals with European ancestry in UK and Ireland, Cardiovascular Dx - AF: Atrial Fibrillation; CAD: Coronary Artery Disease; CT: Coronary Thrombosis; HF: Heart Failure; CF: Coronary Fatality, Comorbidities - OBESE: Obesity; CKD: Chronic Kidney Disease; DIAB: Diabetes; HT: Hypertension; HL: Hyperlipidemia, Cardiovascular Rx - ACGN: Anticoagulant; ARB: Angiotensin Receptor Blocker; ACEIN: ACE Inhibitor; BBL: Beta Blocker; CCBL: Calcium Channel Blocker; FIBR: Fibrate, STAT: Statin; BGN: Biguanide; SUR: Sulphonylurea. The percentage of individuals with incident disease outcome is given atop every bar.

For Rx data, we extracted drugs used for CVD related Dx such as blood clot preventing anticoagulants (ACGN), Angiotensin II Receptor Blockers (ARB) prescribed for HT, blood pressure lowering Angiotensin-converting enzyme inhibitors (ACEIN), and Beta-blockers (BBL). We defined one representative variable for CVD Rx, combining these medication usage indicator variables as CVD Rx = *{*ACGN ∨ ARB ∨ ACEIN ∨ BBL*}*. Similarly for HL, we also defined HL Rx = {STAT ∨ FIBR}, DIAB Rx, for diabetes as DIAB Rx = {BGN ∨ SUR}, where BGN and SUR stands for Biguanides and Sulfonylureas, respectively. HT Rx, for hypertension was defined as presence of Calcium Channel Blockers (CCBL).

### 1.2 Genomic Analyses

We performed genomic quality control (QC) on the imputed imputed genotype data of 104,604 samples and 44 million genetic variants using PLINK [20] and obtained 103,319 samples and

1.73 million variants (more details in Supplementary Note). We performed Principal Component Analysis (PCA) on this data using TeraPCA [21] and obtained the top fifty principal components (PCs).

### 1.3 Association Tests

We performed logistic regression based association tests between CVD Dx and Rx with other demographic and clinical variables including ePRS using statsmodels [22] in Python. Specifically, we identified the association between racial categories and CVD Dx, Rx and related comorbidities.

### 1.4 Feature Selection

To identify informative features and to assess their level of association with different race types we analyzed data for all individuals that had no missing values, a total of 103,986 subjects. We randomly split this data set into 75% (training) and 25% (testing). Race categories included BB, SAS, UKW, EAS, and Mixed. We applied, for each race type, a sub-population-based feature selection method [23] to identify and rank the most informative features. The method was applied in 100 runs for each race on the training set and at the end of the runs all features received a rank between 0 and 100, indicating importance such as higher rank corresponding to higher importance (details in Supplementary Note).

## 2 Results

### 2.1 CVD Dx/Rx and race

The UKB ePRS cohort was used to test for racial bias in CVD related Dx or Rx using clinical variables first. We observe (Figure 2A) that all of the five CVD Dx’s (AF, CAD, CT, CF, and HF) had a protective effect on BB (mean OR=0.63; 95%CI: 0.48-0.87; *p <* 0.01).

**Figure 2:**
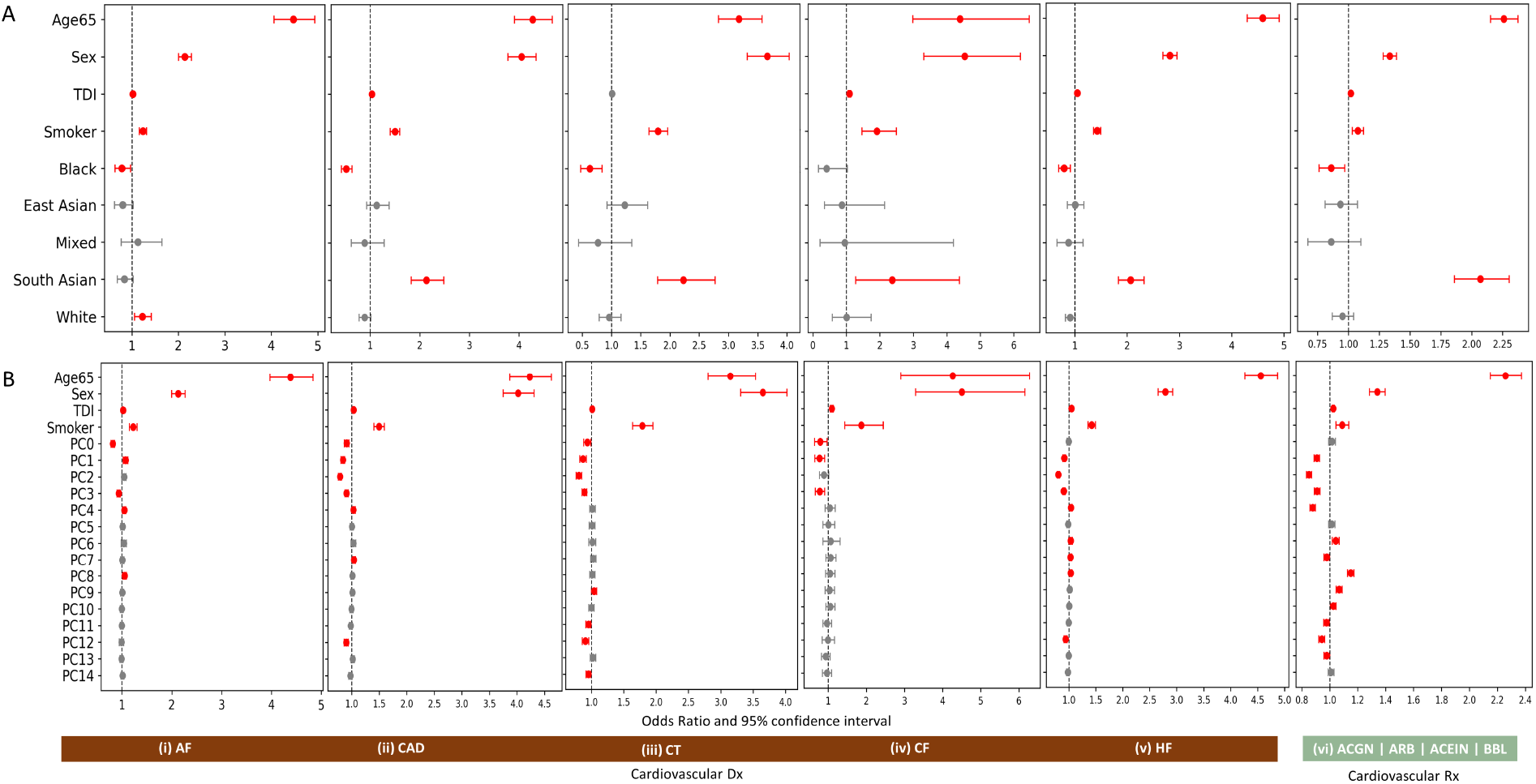
Association between demographic variables such as Age65 (age *>* 65), sex (Male), Townsend Deprivation Index (TDI), smoking status (Smoker), and (A) self-described category of race (B) top 15 PCs and CVD Dx denoted as (i) AF: Atrial Fibrillation (*N* = 4227), (ii) CAD: Coronary Artery Disease (*N* = 4710), (iii) CT: Coronary Thrombosis (*N* = 2316), (iv) CF: Coronary Fatality (*N* = 258), (v) HF: Heart Failure (*N* = 8530), and (vi) Cardiovascular Rx as defined in Methods (*N* = 10636). The significant ORs (*p <* 0.05) are shown in red with error bars reflecting 95% CI.

Individuals with SAS ancestry (Indians, Pakistanis and Bangladeshis) have significantly higher association with severe CVD Dx such as CAD, CT, HF, and even with CF. We observed the same for CVD Rx with a protective effect on BB (OR= 0.72; 95%CI: 0.59-0.89; *p <* 0.0003) and an increased risk in SAS (OR=2.07; 95%CI: 1.87-2.31; *p <* 10^−15^).

One of the early decision points exploring physician bias in CVD involves incident comorbodities of CVD such as HL, HT, and/or diabetes Dx along with their therapeutic interventions. Hence, we sought a test for identifying whether treatment decisions disproportionately impacted BB (Supplementary Figure 2). We found BB to be at a higher risk of diabetes (OR=1.19; 95%CI: 1.07-1.32; *p <* 10^−15^), HT (OR=1.67; 95%CI: 1.54-1.79; *p <* 10^−41^), and obesity (OR=1.32; 95%CI: 1.13-1.55; *p <* 10^−4^), while showing a non-significant effect for HL (OR=0.89; 95%CI: 0.67-1.17; *p <* 0.4). Associations between BB and therapeutics of CVD and related comorbidities followed a similar trend with BB showing a protective effect on lipid-lowering HL Rx such as statins or fibrates (OR=0.77; 95%CI: 0.68-0.87; *p <* 10^−5^) along with a non-significant effect on DIAB Rx (OR=0.96; 95%CI: 0.79-0.1.15; *p <* 0.6) and a strong association with HT Rx (OR=1.35; 95%CI: 1.18-1.54; *p <* 10^−6^). We tested whether the blood lipids levels prior to HL Rx were different among BB vs. the rest of the population and found no significant difference according to prescription for HL patients (OR=1.17; 95%CI: 0.67-1.82; *p <* 0.5), suggesting the differences in Rx rates were not due to observably inconsistent application of clinical practice guidelines (CPGs).

### 2.2 Race and genetics

We sought to identify ancestrally informative genetic features by considering PCA and PRS to see if they may reveal distinct subgroups, and whether identification of subgroups might yield more predictivity of disease and therapy than the ethnographic category of race. We computed PCA on the entire UKB ePRS cohort (Supplementary Figure 3) and chose to keep the top 15 PCs based on the percentage of variance explained by their corresponding eigenvalues (Supplementary Figure 4).

#### 2.2.1 Genetic structure as a proxy for race

We found that the indicator variable of BB had very significantly high OR=6.21 (95%CI: 5.82-6.61; *p <* 0.001) with PC0 (the first PC and so on for the following PCs), followed by PC1 with OR=3.49 (95%CI: 3.21-3.81; *p <* 0.001). They are also significantly associated with some of the other PCs with ORs (albeit with lesser significance) close to one (Figure 3). Similarly, for EAS we observe the highest OR with PC6 with OR=4.39 (95%CI: 3.75-5.13; *p <* 0.001), for SAS we observed association with PC7 and PC8 with mean OR=1.71 (95%CI: 1.55-5.1.89; *p <* 0.001), and for White individuals of European ancestry mostly with PC2 (OR=2.32; 95%CI: 2.15-2.51; *p <* 0.001) and also with PC3 and PC5 (mean OR=1.35; 95%CI: 2.26-1.47; *p <* 0.001). Therefore, we observed all distinct racial categories had significant associations with one or more of the PCs with very high ORs and also with smaller strength of relationship with other PCs. In addition, we found that BB were associated with individuals who are younger than 65 years (OR=0.5; 95%CI: 0.47-0.52; *p <* 0.001, with Age65 variable denoting individuals older than 65 years), significantly associated with TDI (OR=1.28; 95%CI: 1.27-1.28; *p <* 0.001), concordant with the fact that across all ages BB are more deprived compared to other populations (Supplementary Figure 5), and have a protective effect on smoking (OR=0.55; 95%CI: 0.52-0.58; *p <* 0.001).

**Figure 3:**
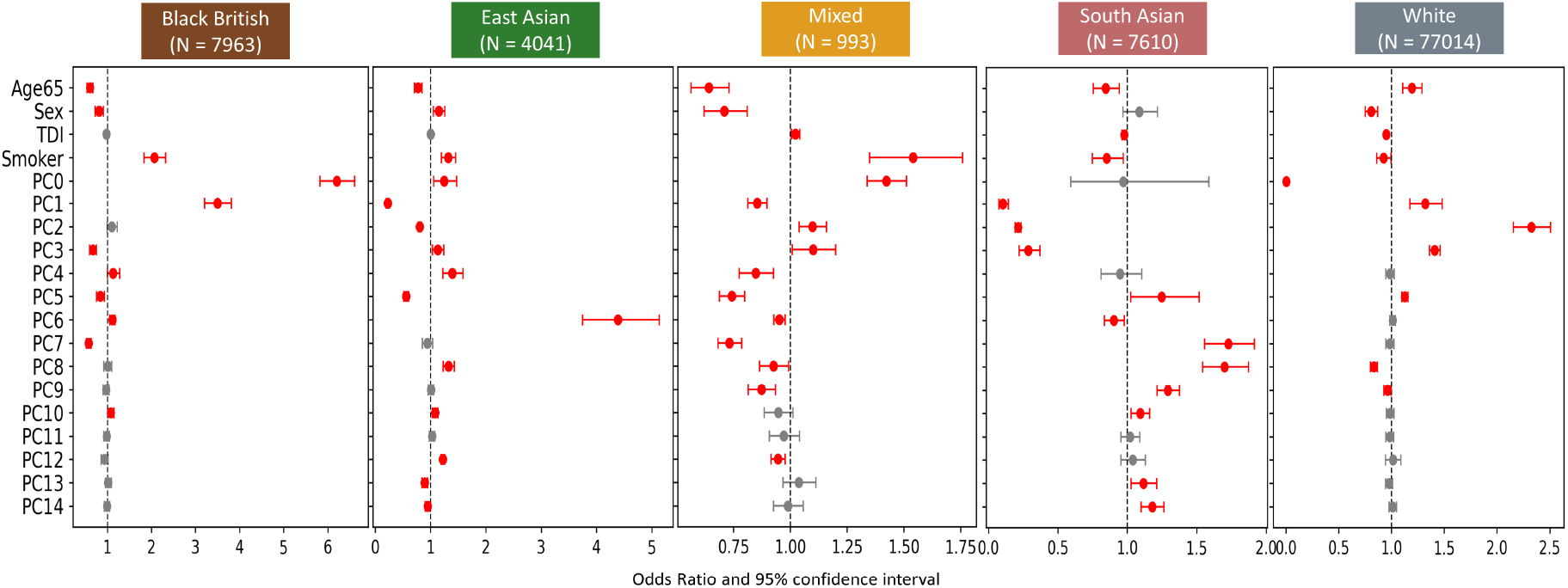
Association between self-described ethnographic category of race and demographic variables which includes the top 15 PCs along with demographic variables such as Age65 (age *>* 65), sex, smoking history and TDI. Each panel corresponds to a race as summarized here: (i) Black British (*N* = 7963), (ii) East Asian (*N* = 4041), (iii) Mixed (*N* = 993), (iv) South Asian (*N* = 7610), and (v) White (*N* = 77014). The significant ORs (*p <* 0.05) are shown in red with error bars reflecting 95% CI.

When we replaced the race variable with genetic structure (PCs) in the association tests between CVD Dx or Rx, we found no substantial change in association of statistics of CVD and associated comorbidities Dx or Rx. For example, when we replace BB with PC0 (associated with highly significant OR=6.21) in the association test, we observed a mean OR=0.89 (95%CI: 0.83-0.95; *p <* 0.002). Although it is above the mean OR of 0.63 with racial categories, the protective effect still remains (Figure 2B; Supplementary Figures 1 and 2).

#### 2.2.2 Race and PRS

CVD and related comorbidities in Dx and Rx showed a protective effect on BB in UKB. Next, we wanted to test whether the predisposed genetics risk of CVD and related traits are associated the race variable, specifically, with BB. We obtained 15 precomputed enhanced PRS (ePRS) from UKB [19] and tested whether the self-described category of race is associated with the ePRS as well as the association of these ePRS stratified by ancestry with respect to Dx. Among the set of ePRS, six were CVD Dx, namely, AF, CVD, CAD, ISS, RHR, and VTE. The rest were ePRS of CVD related comorbidities such as HT, Type I Diabetes (T1D) Type II Diabetes (T2D), BMI, HbA1c, HDL/LDL cholesterol, remnant cholesterol (RMNC), and total cholesterol (TCH).

We observed the racial category of BB was the most associated with the ePRS with the strongest positive association with T2D (OR=1.21; 95%CI: 1.17-1.24; *p <* 10^−36^), followed by RMNC (OR=1.11; 95%CI: 1.07-1.14; *p <* 10^−12^), ISS (OR=1.07; 95%CI: 1.07-1.14; *p <* 10^−10^), CAD (OR=1.11; 95%CI: 1.07-1.14; *p <* 10^−12^), RHR (OR=1.09; 95%CI: 1.06-1.13; *p <* 10^−9^), and other traits. SAS population were the next best predicted by the ePRS with multiple ePRS (CVD, CAD, RHR, VTE, HbA1c, etc.) being strongly associated with a positive effect (Figure 4). This indicates BB and SAS are at most risk of CVD and related diseases.

**Figure 4:**
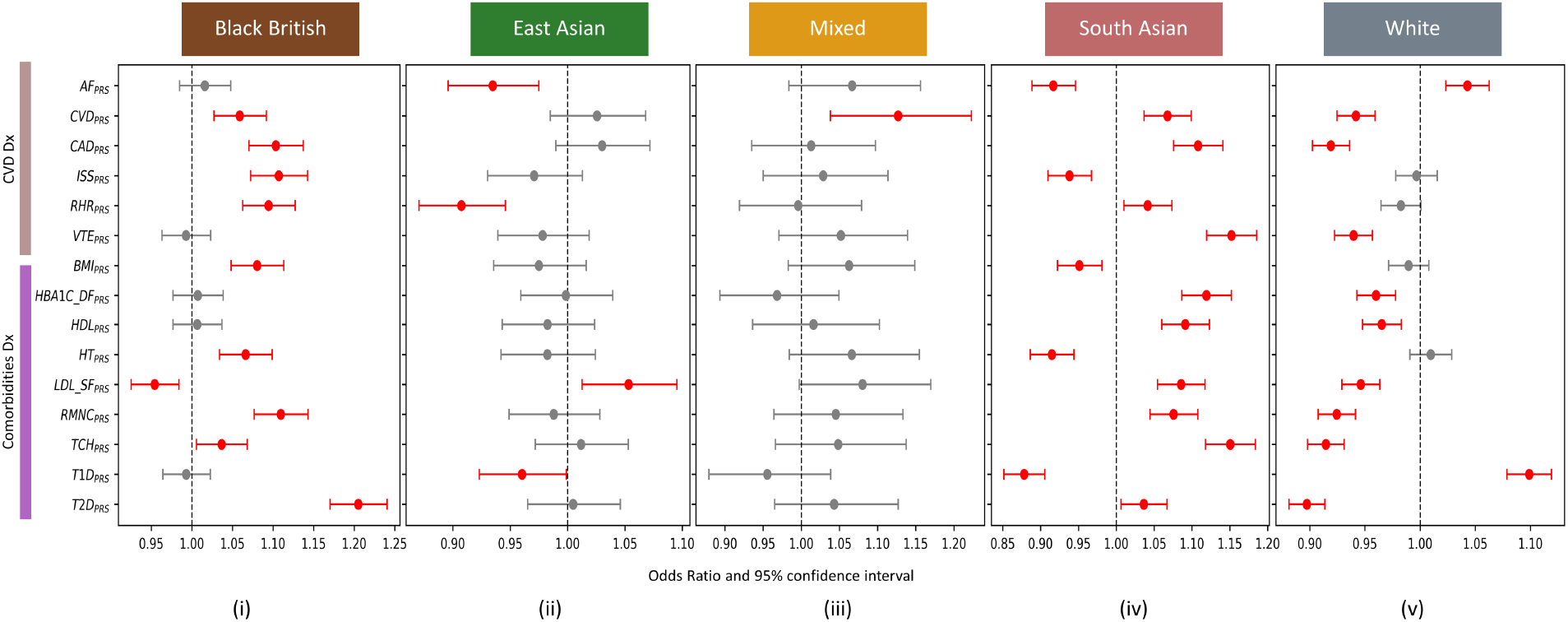
Performance of 15 CVD related ePRS predicting racial categories of (i) Black, (ii) East Asian, (iii) Mixed, (vi) South Asian, and (v) White. The association tests were corrected for age and sex. The significant associations with 95% CI are marked in red.

We also tested the predictive performance for five ePRS, namely, AF, CAD, CVD, HT, and T2D (abbreviated as PRS AF, PRS CAD, PRS CVD, PRS HT, and PRS T2D, respectively) with respect to the Dx, for each population after correcting for age, sex, and top 15 PCs. We observed all of these PRS to be predictive of the Dx significantly and the risk for BB was uniformly present across all these diseases, taking into account transferability of issues of PRS computed from European ancestry individuals to other populations (Supplementary Figure 4).

#### 2.2.3 Predicting race

Predicting BB using the feature selection method we observed the following features received the highest ranks: PRS T2D (98), Age65 (95), PRS AF (94), Smoker (93), and HT (91). Aggregating all of the top features we found the same set of top ten features across all race variables, albeit with varying ranks for each racial category. We found Age65, HT, smoking status, PRS AF, PRS T2D, PRS ISS, DIAB, TDI, PRS LDL SF, and STAT were the top ten features (in decreasing order of importance) predicting race in this data set (Figure 5a).

**Figure 5:**
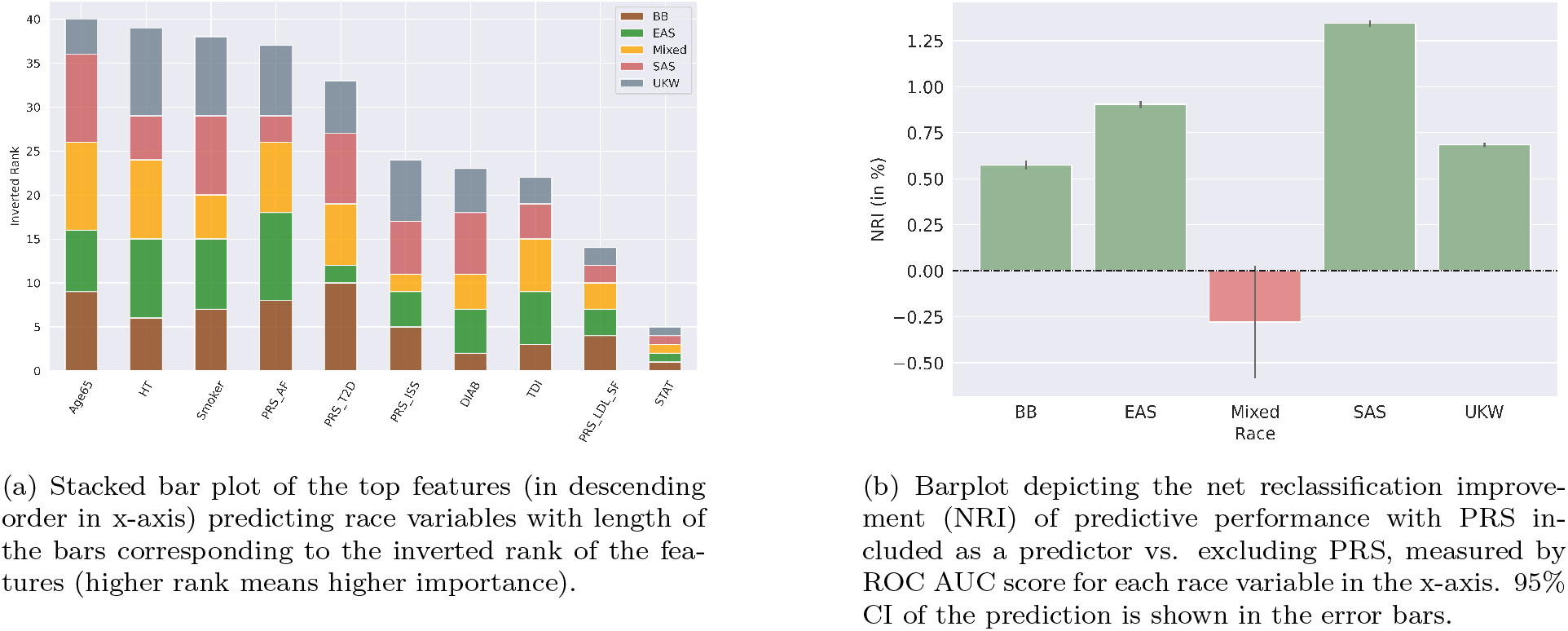
Predictive performance of racial categories.

We sought to use these top features to quantify whether PRS features contribute to the predictivity of a self-described racial category. For each category, we used the testing set to assess prediction performance and applied 100 permutations of randomly selected sub-derivation and sub-validation sets using the testing set. We took the average AUC of these 100 runs to calculate final AUCs and 95 percent confidence intervals, considering all selected features as well as all selected features excluding PRS features. We observed that BB had the highest prediction accuracy of 79.15% (95%CI: 0.77-0.80), followed by SAS (75.28%; 95%CI: 0.73-0.77), UKW (74.74%; 95%CI: 0.73 - 0.75), EAS (63.98%; 95%CI: 0.61-0.66), and Mixed (63.85%; 95%CI: 0.58-0.69) with all features, including PRS features as predictors (Supplementary Figure 7). Furthermore, we observed, that including PRS features as predictors versus excluding them, contributed to positive net reclassification improvement (NRI) (measured as the difference between the predicitivity of the racial category using the two feature sets) for all races except the Mixed category. The maximum positive NRI was observed in SAS, followed by EAS, UKW, and BB (Figure 5b).

## 3 Discussion

Testing for clinical bias in treating BB and other underrepresented participants in the UKB poses multiple problems. First, the population’s age structure is younger, and older age is a primary associating variable with CAD and related CVD Dx. This was observed in our cohort as BB tend to have lower odds of being older than 65 compared to the population (Figure 3). Second, some CPGs are specific in treatment modalities, such as application of CCBL for HT. Two opportunities of identifying bias are (1) whether the pathway from HL through CF shows difference in rates of Dx between clinical screening and treatment versus the British death registry, and, (2) whether patients identified with HL are treated at different rates with lipid management Rx based on racial identification. A second question to identify bias is whether racial identification may under-diagnose disease compared to genetics. Again, there are two opportunities to investigate this (1) by use of PCs as ancestrally informative genetic measures used instead of an indicator variable of race, and (2) using PRS as a predisposed measure of genetic risk of CVD and comparing it with observed CVD Rx and Dx for each racial category.

Investigating the association of race with HL Dx and Rx, we observed no significant change in the direction of effect, with both HL Dx and Rx showing a protective effect for BB, after controlling for age, sex, TDI, and smoking status. Exploring the interaction of age with BB and its association with HL, we found that if the patient is 65 years or younger and BB, it is significantly protective. If the patient is BB and over 65 years, the OR = 1.69 (95%CI: 0.927-3.1; *p* = 0.082), hence statistical significance is insufficient to resolve whether BB is relatively protective against age greater than 65. Although, BB was not significantly associated with HL (Supplementary Figure 1), it showed a significant positive association with HT and obesity which are notable antecedents of CVD Dx [24]. We observed, HL Rx also showed a protective effect on BB (Supplementary Figure 2) and similarly, following the pattern in Dx variables,

HT Rx, showed a positive association with BB and DIAB Rx had no statistical significance while associated with BB. We note, that the low sample sizes for both DIAB Rx (n=2710) and HL Dx (n=1636) with BB (n=3107) with respect to rest of the population (n=100,212) may play a role for the lack of statistical significance for these associations. Reported diagnoses show higher risk of DIAB and HT indicates first that screening is being applied, and that, while accessibility and trust may reduce BB participation, the risk among BB who do get checkups are being tested. Therefore, it does not account for lower rates of Dx of HL, and CVD Dx among BB, especially given higher odds of HT and DIAB. Since autopsy diagnoses, as independent measures of disease than clinical physician visits, show similar levels of CVD Dx involvement, the likelihood that the result is due to physicians failing to diagnose these conditions is reduced. Further, even though the likelihood that a physician might prescribe a HL Rx given a HL Dx, direct comparisons of LDL levels between groups resulting in HL Rx therapies suggests guidelines are not being differentially applied.

Racial categorization is poorly defined, driven by historical interactions, and loaded with false assumptions [4]. There is a significant possibility that such classifications poorly align with ancestrally informative mutations (AIMs) relevant to pathogenic processes, and to unfair diagnoses and prescribed therapies. However, presumed race has impacted enfranchisement for economic and cultural opportunity resulting in deprivation with medical impact that may spuriously correlate with AIMs, impacting people with, various ethnicities [25]. It is possible that the long British colonial history may have brought a very diverse range of distinct populations which are now labeled simply as Black British. Therefore, racial classification may hide genetically distinctive groups with differential response to diseases. PCA applied to genome-wide single nucleotide variants is strongly reflective of ancestry [26]. In populations structured by ethnicity, these genetic characteristics may include variants that may reflect linkage that are physiologically relevant, but they may also include diet, or economic status correlations [27]. PCA will provide more information about cryptic admixtures that may also impact associations with diseases. Therefore, we sought to identify whether there was an ancestrally genetic community labeled as BB or admixed that may show different risks of CVD, and which might point towards therapies that would save more lives. Apart from a continuum of admixture, no distinctive ancestral group was identifiable. PCs were very strongly predictive of ethnographic classifications. The PCs captured similar predictive information for CVD Dx, Rx, and comorbidities that the social construct “Black British” identifies. BB tended to be protective of incident CVD Dx and Rx (Figure 2) and the same was reflected when PCs were used instead of racial categories.

PRS is a single-score summary of an individual’s pre-disposed genetic risk or liability of a trait or disease. PRS in UKB has revealed 20-fold more people of greater risk of CVD Dx than previous studies [28]. BB and SAS were the populations with most positive associations across ePRS of CVD and comorbidities Dx obtained from UKB (Figure 4). PRS for CAD has been shown to be at higher odds for SAS populations in prior work [29] and here, we reveal the first known systematic evaluation for BB populations who are shown to be at higher odds of PRS for CVD and related comorbid traits. The ethnographic category of BB was also predicted by PRS of AF and T2D with high accuracy, along with other demographic variables (Figure 5a). Thus, although BB had a protective effect for CVD Dx and Rx, we show that the population has a higher odds of the pre-disposed genetic risk of CVD and its comorbidities. Therefore, we posit that instead of this higher risk of CVD, BB populations either do not get diagnosed or treated for CVD. This observation is also influenced by the lack of enrollment of BB individuals in UKB and other large biobanks. We note that PRS has shown to suffer from lack of transferability and bias across populations [13], primarily due to overwhelming presence of European ancestry individuals in large biobanks. This phenomenon was mitigated during the release of ePRS in UKB [19]. Recent advances in applications of machine learning also help establish more generalizable PRS for clinical use [30,31]. This calls for more diversity in enrollment and evaluation of underrepresented populations for creating a more equitable healthcare system.

## 4 Conclusion

We investigated for bias in Dx and Rx for CVD and related metabolic diseases by associating them with the self-described ethnographic category of race in a large biobank. We found that the OR for Dx and Rx for these diseases did not vary for the racial categories. The OR for dyslipidemia Dx and CT were proportional to the OR for death due to MI in the death registry, which offered an independent diagnostic review. Ancestral genetic analysis did not identify a distinctive group of BB that might have carried a higher rate of disease or mortality, and the genetic components associated with BB identifications was similarly predictive of relevant metabolic syndrome and CVD Dx. Interestingly, we observed among hyperlipidemics, being BB tended to suppress prescription, even among older patients. except among those diagnosed as obese, in which case, prescription was more likely. PRS analyses, showed that BB and SAS were of higher risk of PRS for CVD and metabolic syndrome Dx, however, the Dx and Rx rates were negatively associated with BB. This indicates that although racial disparities exist in terms of pre-disposed genetic risk of CVD Dx, it is not reflected in Dx and Rx rates in large biobanks. Thus, a more concerted effort is required to study populations with African, Asian and other underrepresented ancestries to create a fair healthcare system.

## Data Availability

All data is available from UK Biobank upon request.

## 5 List of Abbreviations

CVD: Cardiovascular diseases
UKB: UK Biobank
OR: Odds Ratio
AF: Atrial Fibrillation
CAD: Coronary Artery Disease
PRS: Polygenic Risk Scores
AHA: American Heart Association
Dx: Diagnoses
Rx: Therapeutics
ACE: Angiotensin Converting Enzyme
PCs: Principal Components
BB: Black British
CT: Coronary Thrombosis
CF: Coronary Fatality
HF: Heart Failure
OBESE: Obesity
CKD: Chronic Kidney Disease
DIAB: Diabetes
HL: Hyperlipidemia
HT: Hypertension
EAS: East Asians
SAS: South Asians
UKW: UK White
ISS: Ischemic Stroke
RHR: Resting Heart Rate
VTE: Venous Thromboembolic Disease
BMI: Body Mass Index
LDL: Low Density Lipoprotein
HDL: High Density Lipoprotein
STAT: Statins
FIBR: Fibrates
ACGN: Anticoagulants
ARB: Angiotensin II Receptor Blockers
ACEIN: Angiotensin Converting Enzyme Inhibitors
BBL: Beta Blockers
CCBL: Calcium Channel Blockers
PCA: Principal Component Analysis
Age65: Age above 65 years

## 6 Declarations

### 6.1 Ethics approval and consent to participate

Data analysis was performed under UK Biobank application 50658 using existing publicly available and deidentified data and was IRB exempt.

### 6.2 Consent for publication

Not applicable

### 6.3 Availability of data and materials

The feature selection tool used in this study is available in https://github.com/IBM/spbfs

### 6.6 Authors’ contributions

AB and DEP conceived the project. AB and DEP performed epidemiological analyses. AB performed genomic analyses. UK performed the race prediction task. AB, DEP, and UK wrote the manuscript. KN and LP participated in discussions, reviewed, and supervised the project.

## 6.7 Acknowledgements

Not applicable

