## Supplementary Information for "Role of genetics in capturing racial disparities in cardiovascular disease"

### Supplementary Materials

#### Methods

##### Quality Control

We performed genomic quality control (QC) on the imputed genotype data of 104,604 samples and 44 million high quality imputed variants (INFO $> 0.3$) using PLINK [20]. We removed missing samples and variants with 2% missing values, minor allele frequency (MAF) $< 0.05$, Hardy-Weinberg Equilibrium (HWE) $< 10^{-6}$ and removed samples with gender discrepancy, more than three standard deviations in heterozygosity rates along with closely related individuals ($\hat{\pi} > 0.125$). We finally obtained 103,319 samples and 1.73 million variants after QC.

##### Principal Component Analysis

We performed Principal Component Analysis (PCA) on the QC data using `TeraPCA` [21] and obtained the top fifty principal components (PCs).

##### Feature Selection

To identify informative features and to assess their level of association with different race types we analyzed data for all individuals that had no missing values, a total of 103,986 subjects. We randomly split this data set into 75% (training) and 25% (testing). Race categories included BB, SAS, UKW, EAS, and Mixed. We applied, for each race type, a sub-population-based feature selection method [23] to identify and rank the most informative features. The method was applied in 100 runs for each race on the training set and at the end of the runs all features received a rank between 0 and 100, indicating importance such as higher rank corresponding to higher importance.

We obtained the rank for each feature corresponding to each racial category after applying the feature selection method and ordering them from 1 to 10 with respect to the rank. We term this as the original rank. Thereafter, as we observed there were ten features with the highest original rank across all race categories, we obtained the inverted rank of these features relative to each racial category. We define the inverted rank as the following

$$\text{Inverted Rank} = 10 - \text{Original Rank}$$

The inverted rank was reported in Figure 6). This was done for visualization purposes as the stacked bars reflect the importance of each feature to each racial category. A larger bar corresponding to high importance.

### Results

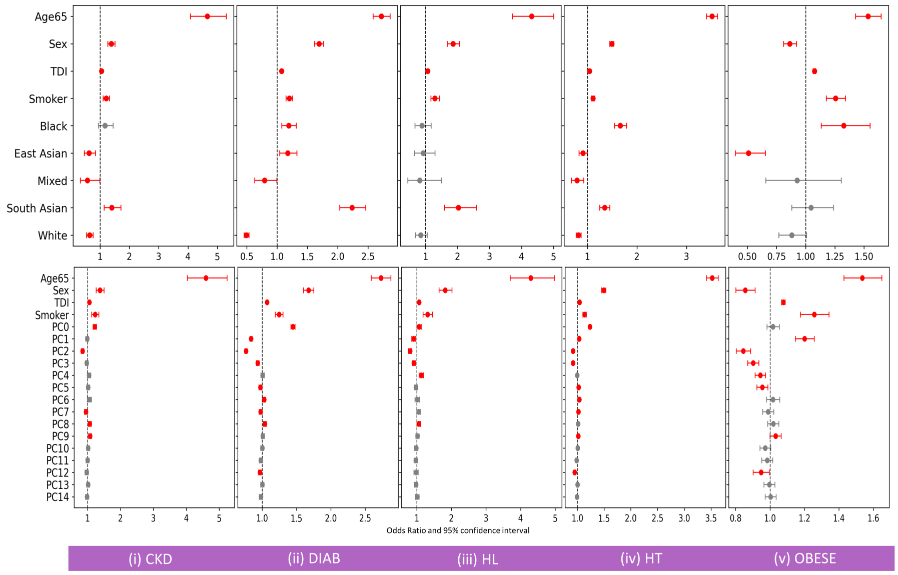


Figure 1: Association between demographic variables such as Age65 (age $> 65$), sex (Male), Townsend Deprivation Index (TDI), smoking status (Smoker), and (A) self-described category of race (B) top 15 PCs and diagnoses of CVD related comorbodities such as (i) CKD: Chronic Kidney Disease ($N = 2165$), (ii) HL: Hyperlipidemia ($N = 1636$), (iii) HT: Hypertension ($N = 34817$), (iv) OBESE: Obesity ($N = 4020$). The significant ORs ($p < 0.05$) are shown in red with error bars reflecting 95% CI.

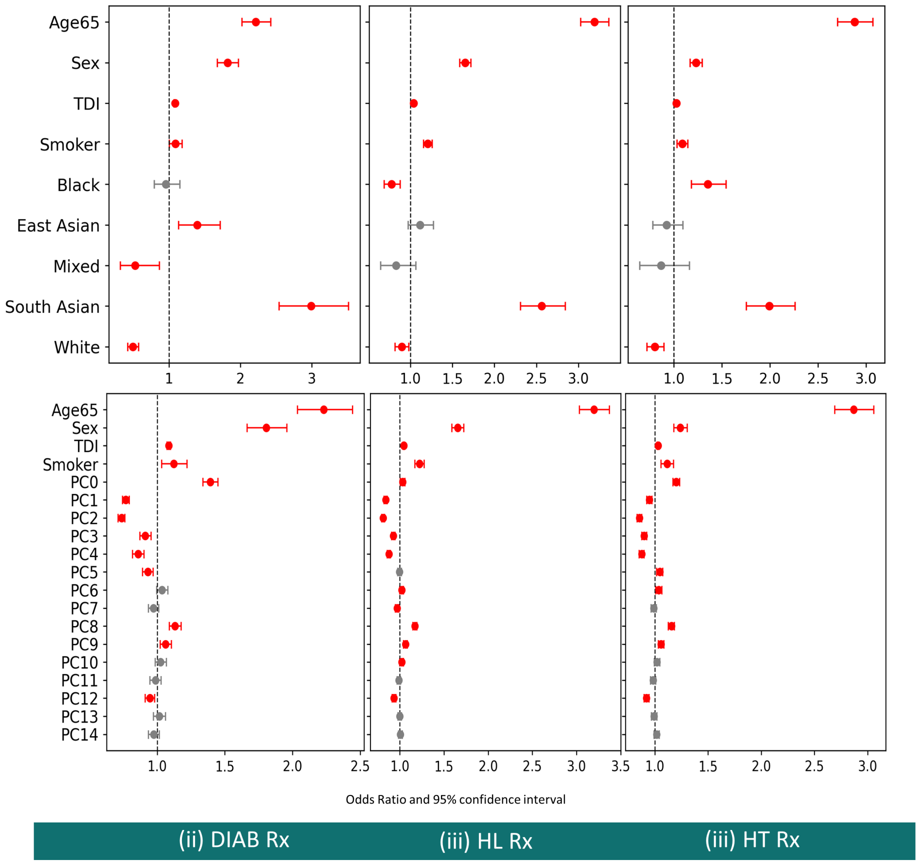


Figure 2: Association between demographic variables such as Age65 (age $> 65$), sex (Male), Townsend Deprivation Index (TDI), smoking status (Smoker), and (A) self-described category of race (B) top 15 PCs and diagnoses of CVD related comorbodities such as (i) DIAB Rx: Diabetes Rx ($N = 2710$), (ii) HL Rx: Hyperlipidemia Rx ($N = 10868$), (iii) HT Rx: Hypertension Rx ($N = 6805$). The Rx variables are as defined in Methods. The significant ORs ($p < 0.05$) are shown in red with error bars reflecting 95% CI.

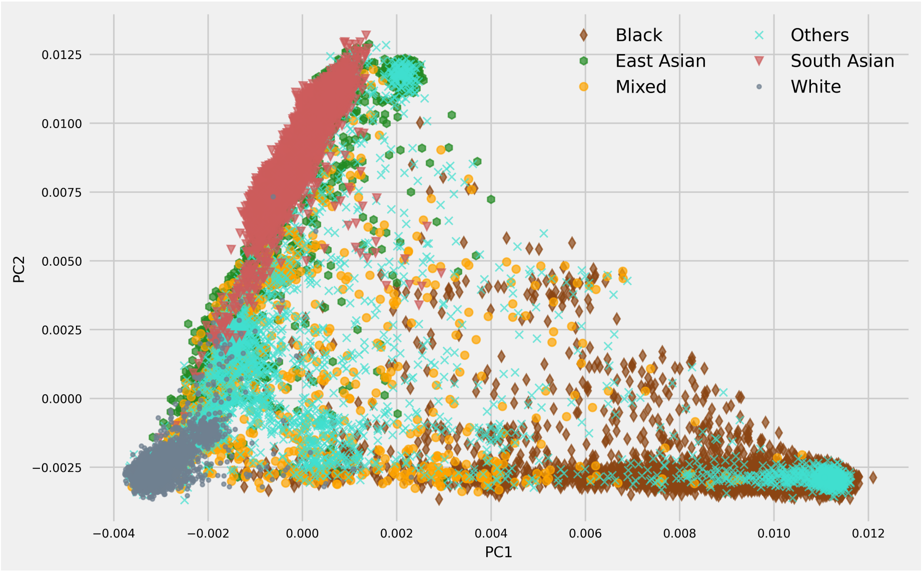


Figure 3: Top two PCs from the PCA of the UKB ePRS with 103,319 individuals after quality control.

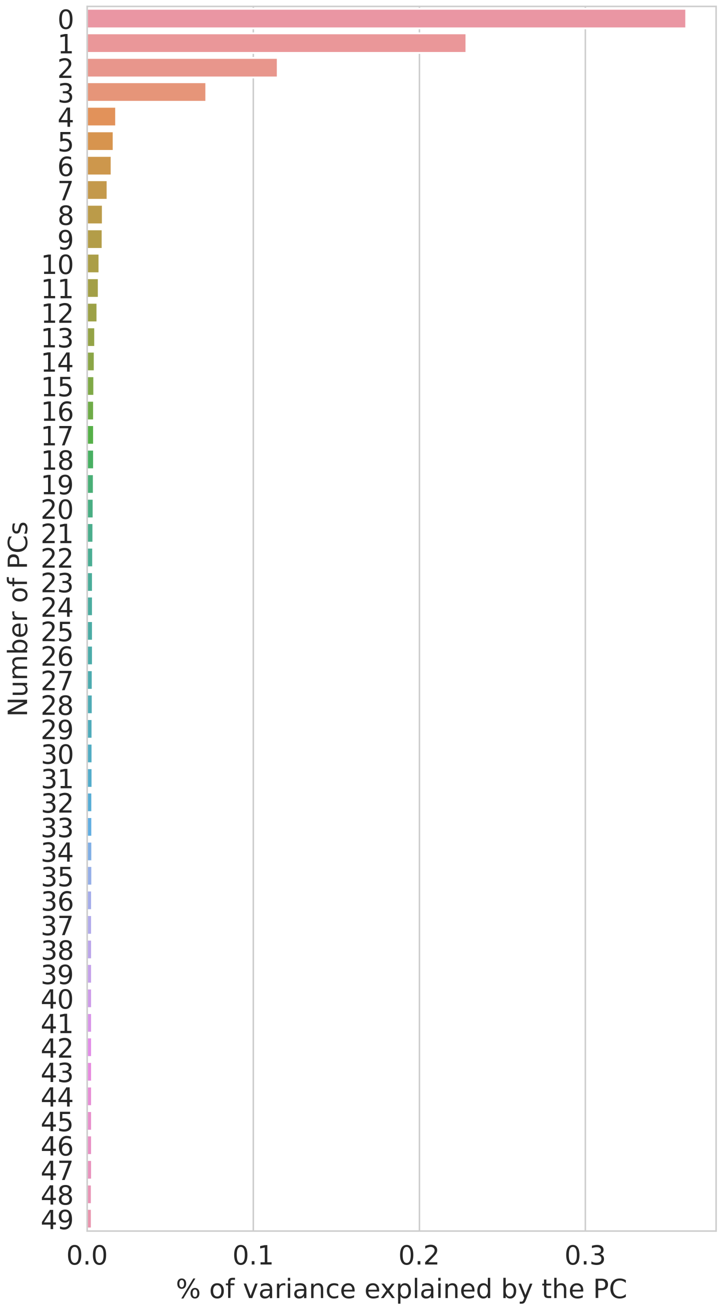


Figure 4: Percentage of variance explained by the PCs.

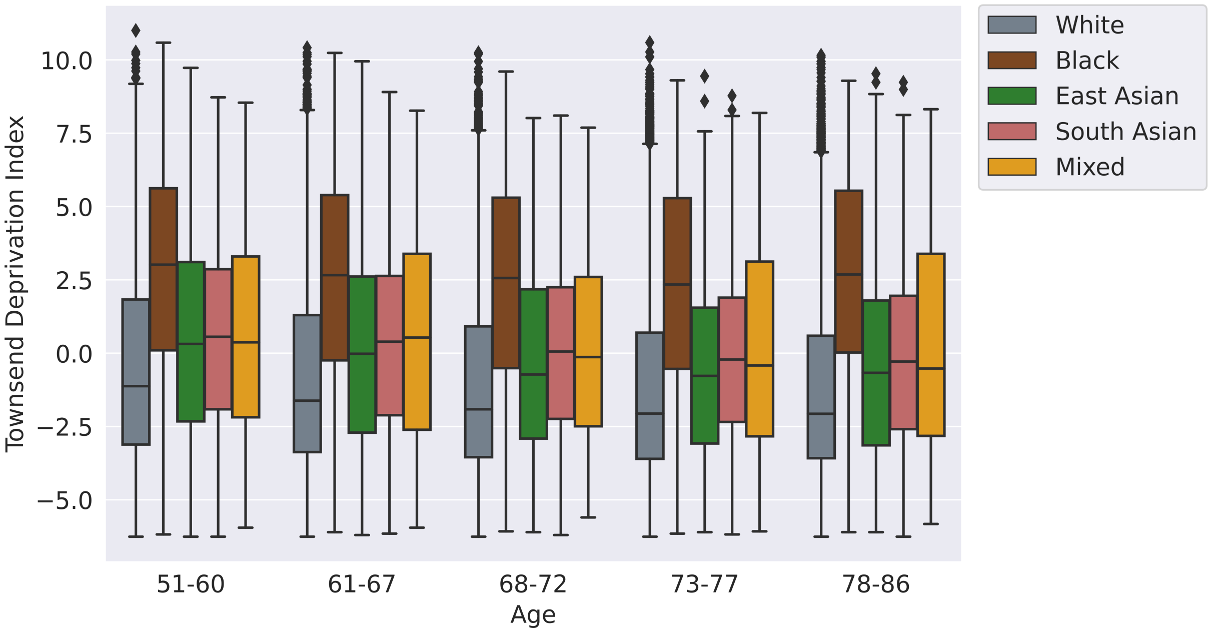


Figure 5: Box and whisker plot denoting the relationship with every racial category with Townsend Deprivation Index, stratified by age.

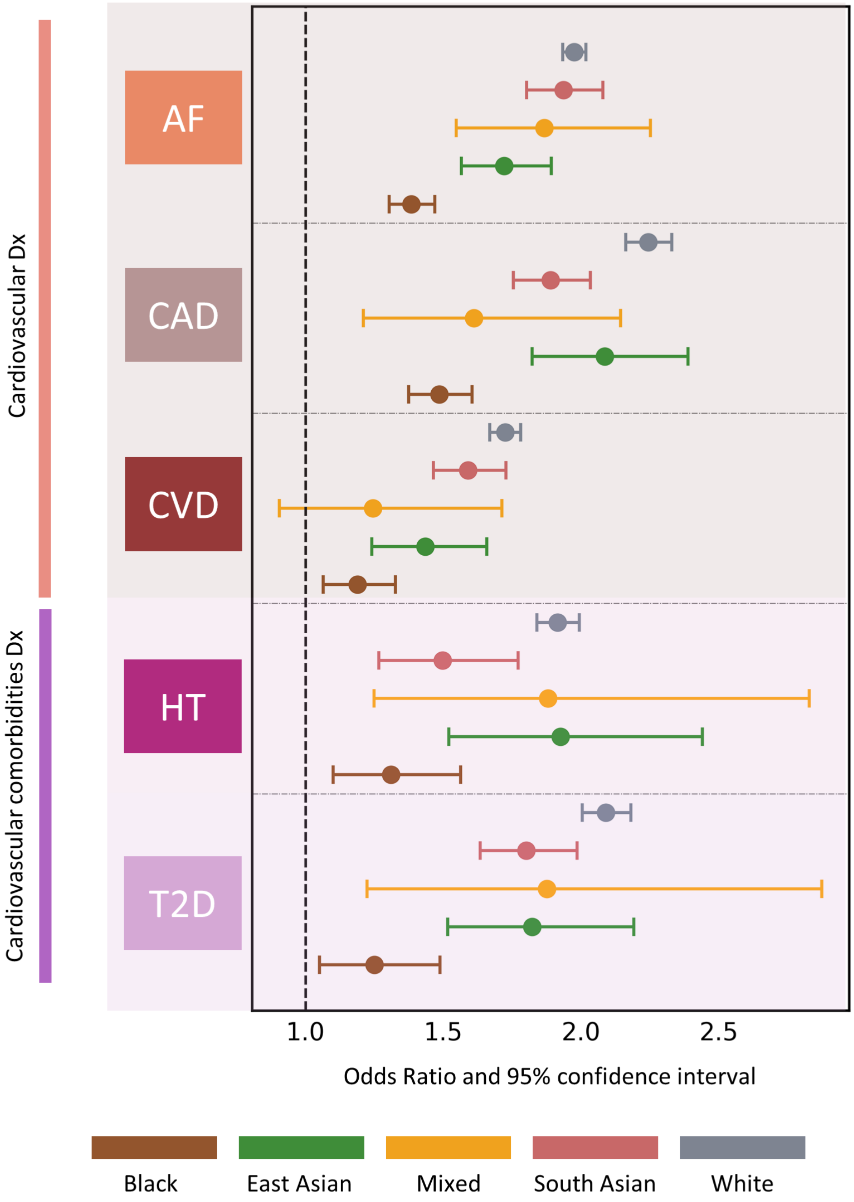


Figure 6: Predictive performance of five ePRS with respect to their Dx, stratified by racial categories and corrected for age, sex, and top 15 PCs.

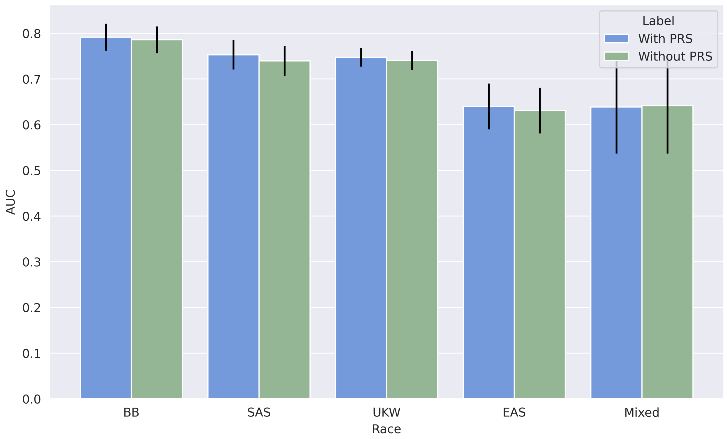


Figure 7: Predictive performance (as measured by AUC) of the racial categories with and without including PRS as predictors.
